# Comparative performance of COVID-19 Test Methods in Healthcare Workers during the Omicron Wave

**DOI:** 10.1101/2023.09.25.23296099

**Authors:** Emma Tornberg, Alexander Tomlinson, Nicholas T.T. Oshiro, Esraa Derfalie, Marcel E. Curlin

## Abstract

**Introduction:** The COVID-19 pandemic presents unique requirements for accessible, reliable testing, and many testing platforms and sampling techniques have been developed. However, not all test methods have been systematically compared to each other or a common gold standard, and the performance of tests developed in the early epidemic have not been consistently re-evaluated in the context of newly emerging SARS-CoV-2 variants.

**Methods:** We conducted a repeated measures study with adult healthcare workers presenting for SARS-CoV-2 testing. Participants were tested using seven test modalities: PCR with samples from the nasopharynx, oropharynx, and saliva; and BinaxNOW and iHealth antigen–based rapid detection tests (AgRDT) sampling the oropharynx and the nares. Test sensitivity was compared using any positive PCR test as the gold standard.

**Results:** 325 individuals participated in the study. PCR tests were the most sensitive with saliva PCR at 0.957 ± 0.048, nasopharyngeal PCR at 0.877 ± 0.075, and oropharyngeal PCR at 0.849 ± 0.082. Standard nasal rapid antigen tests were less sensitive but roughly equivalent at 0.613 ± 0.110 for BinaxNOW brand and 0.627 ± 0.109 for iHealth. Oropharyngeal rapid antigen tests were the least sensitive with BinaxNOW and iHealth brands at 0.400 ± 0.111 and 0.311 ± 0.105 respectively.

**Conclusion:** PCR remains the most sensitive testing modality for COVID-19, with saliva PCR being significantly more sensitive than oropharyngeal PCR and equivalent to nasopharyngeal PCR. Saliva testing has patient comfort and financial benefits, making it a preferred testing modality. Nasal AgRDTs are less sensitive than PCR though more accessible and convenient.

## Introduction

The dramatic appearance of the highly transmissible SARS-CoV-2 pandemic on the world stage in late 2019 created an urgent need for rapid, point-of-care diagnostic tests that could accurately detect infectious cases. By 2022, a wide range of testing modalities had been developed, generally relying on either nucleic acid amplification (NAAT) or detection of viral antigens by lateral flow chromatography (antigen rapid diagnostic tests, or Ag-RDTs). NAAT-based tests are generally regarded as the most sensitive testing modality but require laboratory processing, have longer result turnaround times, and may yield false positive results in those with recent resolved infection. Use of at-home Ag-RDTs has been widespread, with 20% of individuals reporting use of one of these tests during the last 30 days during the period of omicron variant dominance (omicron wave)^1^. Pre-omicron data showed sensitivity of a single Ag-RDT to be approximately 70%, with positive correlation between higher viral load as estimated by Ct values and sensitivity^2–4^. Early studies during the omicron wave show varied sensitivity between Ag-RDT brands^5^, with some such as the BinaxNOW showing maintained sensitivity of around 65%^6^. Other brands such as iHealth have shown similar sensitivity in detecting delta and omicron variants in the laboratory but have not been as widely validated in epidemiologic studies during the delta and omicron waves^7^.

Several additional factors may affect this performance. Many of these tests were developed and validated before the emergence of B.1.1.529 (omicron) variants of concern and subsequent viral strains^8^. The rapidly evolving nature of variants and subvariants of SARS-CoV-2 lead to questions regarding the validity of previously developed tests with virus changes that may affect viral load or optimal test sample source. Site of testing is another important variable. For NAATs, the most common of which use polymerase chain reaction (PCR) technology, a variety of sample sources have been evaluated including nasopharyngeal swabs (NP), oropharyngeal swabs (OP), and saliva samples. Pre-omicron meta-analyses showed either equivalence in saliva and NP testing^9^, or slightly increased sensitivity of NP testing compared with saliva^10–12^. However, some studies involving early omicron strains showed better sensitivity with saliva swabs compared to mid-turbinate swabs^13,14^. NP testing is more costly than saliva and can be associated with significant discomfort.^15^ The standard collection site for AgRDTs is the bilateral nares^16,17^, but the evidence of better PCR sensitivity in saliva compared to NP raises the question of whether OP AgRDT testing could provide equal or better sensitivity. Given the many available testing platforms and testing sites, lack of standardization between methods, and significant issues relating to cost, performance, and patient experience, we performed a repeated– measures observational study in a sample of adult healthcare workers presenting consecutively for SARS-CoV-2 testing at the Oregon Health and Sciences University Occupational Health clinic.

## Methods

*Regulatory approval*: This study was performed with informed consent from all participating individuals, with approval and regulatory oversight by the Oregon Health and Sciences University institutional review board.

*Recruitment*: We designed a repeated measures study evaluating PCR tests, iHealth Ag-RDTs, and BinaxNOW Ag-RDTs at the OHSU Occupational Health Testing Site. Between January 25, 2022 and March 4, 2022, individuals presenting for SARS-CoV-2 testing for any reason at OHSU Occupational Health were approached consecutively and offered screening for enrollment on a first-come-first-serve basis. Individuals who had tested positive for COVID-19 in the last 90 days were excluded due to risk of false-positive results, and participants were required to be over the age of 18. Following consent, we tested each participant using 7 testing modalities as follows: PCR (nasopharyngeal swab, oropharyngeal swab, saliva sample); BinaxNOW Ag-RDT (nasal swab, oropharyngeal swab); and IHealth Ag-RDT (nasal swab, oropharyngeal swab).

*PCR testing*: Researchers collected NP and OP swabs from participants in accordance with CDC guidelines^18^. For saliva testing, participants were required to not eat or drink anything for 30 minutes prior to sample collection. They were then directed to spit the saliva that pooled in the bottom of their mouth into the collection tube until it reached the volume specified by the manufacturer. Saliva samples were stored on ice and all samples were transported to the OHSU Molecular Microbiology Laboratory for testing on the day of collection. Viral RNA was extracted using the King Fisher MagMAX Viral/Pathogen Nucleic Acid Isolation Kit, then samples were tested using Taqpath™ COVID-19 Multiplex RT-PCR which targets the Covid-19 S-gene, N-gene, and ORF1ab. A positive test result required positive readings in 2/3 targets at a cycle threshold <40.

*AgRDT testing*: Researchers collected and ran nasal swabs in accordance with manufacturer specifications^16,17^, except that samples were collected and tested outdoors at ambient temperatures ranging from 40-60°F (manufacturer suggest testing at “room temperature”). OP Ag-RDT samples were collected in accordance with CDC OP guidelines and samples were run otherwise in accordance with manufacturer guidelines. All samples were read out after the recommended run time by the collecting researcher.

Statistical Analysis: Symptom data, test results, and sequencing data were described using frequencies and percentages. Likelihood ratios for symptoms and pairs of symptoms were calculated from contingency tables. Using a positive test on any PCR test as the gold standard, contingency tables were generated for each testing modality, and sensitivities and specificities were calculated. McNemar’s test was used for P-values. A P-value of <0.05 was considered statistically significant. Statistical analyses were conducted using Python version 3.10.8.

*Data agreement:* all deidentified data will be made publicly available on request by writing to.

## Results

*Patients and samples*: 325 individuals participated in this study. Six were not tested with BinaxNOW kits due to supply shortages, one did not receive OP iHealth testing due to user error, and 32 were not tested in saliva due to eating or drinking within 30 minutes of the time of testing. All others were tested using all testing modalities. 75 individuals were positive on at least one test; 8 individuals were positive on a single test; 19 individuals were positive on 1 or more PCR tests but negative on all antigen rapid detection tests; 0 were negative on all PCR tests but positive on one or more AgDT; and 13 individuals were positive on all tests performed. Distribution of number of positive tests per individual and types of positive tests per number of positive test are shown in figures 1 and 2.

**Figure 1:**
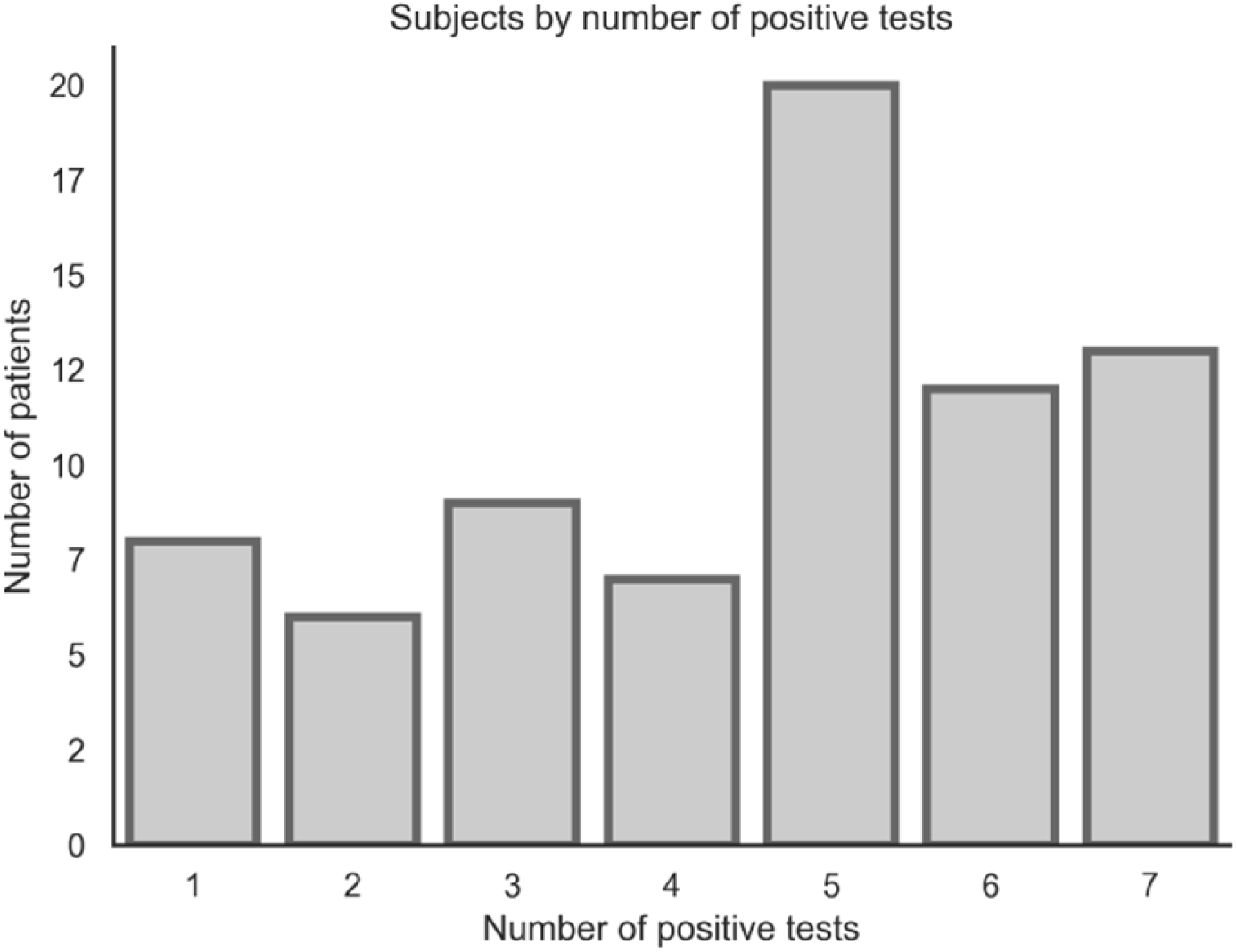
Subjects by number of positive tests.

*Symptoms*: 27 different symptoms were reported across all (A) participants. 16 of these were reported in individuals that tested positive (P) (table 1). Sore throat (A = 23.8%, P = 21.3%), cough (A = 12.6, P = 20.0), and headache (A = 11.8, P = 11.1) were the most commonly reported symptoms for both A and P participants. Likelihood ratios (LR) for single symptoms ranged from 0.00 (nausea) to 3.33 (fever) (Table 2). Likelihood ratios of a positive test were highest for the combinations of chills and cough (26.7), fever and headache (5.3), and headache and cough (5.2) (Table 3).

**Table 1:** Symptoms Reported by All/Positive participants. Total number of participants reporting each symptom is reported on the left side of each pair, with percentage in that category listed on the right.

| Symptom | All Participants |  | Positive Participants |  |
| --- | --- | --- | --- | --- |
|  | Number reporting | Proportion reporting | Number reporting | Proportion reporting |
| sore throat | 165 | 0.2381 | 46 | 0.2130 |
| cough | 87 | 0.1255 | 43 | 0.1991 |
| headache | 82 | 0.1183 | 24 | 0.1111 |
| congestion | 74 | 0.1068 | 18 | 0.0833 |
| fatigue | 60 | 0.0866 | 12 | 0.0556 |
| body aches | 60 | 0.0866 | 23 | 0.1065 |
| rhinorrhea | 43 | 0.0620 | 8 | 0.0370 |
| fever | 36 | 0.0519 | 18 | 0.0833 |
| chills | 21 | 0.0303 | 10 | 0.0463 |
| nausea | 12 | 0.0173 |  |  |
| diarrhea | 9 | 0.0130 |  |  |
| shortness of breath | 6 | 0.0087 | 3 | 0.0139 |
| chest tightness | 5 | 0.0072 | 2 | 0.0093 |
| dizziness | 5 | 0.0072 | 2 | 0.0093 |
| sneezing | 4 | 0.0058 | 1 | 0.0046 |
| loss of smell | 4 | 0.0058 | 2 | 0.0093 |
| vomiting | 3 | 0.0043 |  |  |
| ear pain | 3 | 0.0043 | 2 | 0.0093 |
| sinus pressure | 3 | 0.0043 | 1 | 0.0046 |
| hot flashes | 2 | 0.0029 |  |  |
| stuffy nose | 2 | 0.0029 |  |  |
| gastrointestinal | 2 | 0.0029 |  |  |
| neck stiffness | 1 | 0.0014 |  |  |
| loss of taste | 1 | 0.0014 |  |  |
| night sweats | 1 | 0.0014 | 1 | 0.0046 |
| loss of appetite | 1 | 0.0014 |  |  |
| insomnia | 1 | 0.0014 |  |  |

**Table 2:** Likelihood ratio of a positive vs a negative test.

| Symptom | Likelihood ratio |
| --- | --- |
| sore throat | 1.289 |
| cough | 3.258 |
| headache | 1.379 |
| congestion | 1.071 |
| fatigue | 0.833 |
| body aches | 2.037 |
| runny nose | 0.762 |
| fever | 3.333 |
| chills | 3.030 |
| nausea | 0.000 |

**Table 3:** Likelihood Ratios for Positive Tests of Paired Symptoms.

|  | sore throat | cough | headache | congestion | fatigue | body aches | runny nose | fever | chills | gi | nausea |
| --- | --- | --- | --- | --- | --- | --- | --- | --- | --- | --- | --- |
| sore throat |  |  |  |  |  |  |  |  |  |  |  |
| cough | 3.60 |  |  |  |  |  |  |  |  |  |  |
| headache | 1.35 | 5.15 |  |  |  |  |  |  |  |  |  |
| congestion | 0.93 | 3.33 | 1.78 |  |  |  |  |  |  |  |  |
| fatigue | 0.77 | 2.14 | 1.11 | 0.63 |  |  |  |  |  |  |  |
| body aches | 2.63 | 3.94 | 2.04 | 0.71 | 0.71 |  |  |  |  |  |  |
| runny nose | 0.53 | 0.95 | 0.37 | 0.00 | 1.67 | 0.95 |  |  |  |  |  |
| fever | 4.07 | 5.00 | 5.33 | 2.22 | 0.67 | 2.50 | 0.00 |  |  |  |  |
| chills | 2.78 | 26.67 | 3.33 | 2.00 | 0.56 | 2.67 | 3.33 | 4.44 |  |  |  |
| gi | 0.00 | 0.00 | 0.00 | 0.00 | 0.00 | 0.00 | 0.00 | 0.00 |  |  |  |
| nausea | 0.00 | 0.00 | 0.00 | 0.00 | 0.00 | 0.00 | 0.00 | 0.00 |  | 0.00 |  |

*Test Sensitivity and Specificity:* Test sensitivity ranged from 0.311 ± 0.105 for OP iHealth to 0.957 ± 0.048 for saliva PCR. Test specificity was 1 (Table 4) for all tests examined. Among individuals positive by any PCR, 68.0% were positive on all three PCR tests (Figure 3) and among those positive for one or more PCR tests, 74.7% had at least one positive AgRDT. Individuals positive on any PCR were positive on at least one nasal AgRDT and at least one OP AgRDT 34.7% of the time, positive on at least one nasal AgRDT but no OP AgRDTs 28.0% of the time, and positive on at least one OP AgRDT but no nasal AgRDTs 12.0% of the time (Figure 4). Sensitivities of PCR tests were significantly higher than AgRDTs (all p<0.0001). There were no significant differences in performance between BinaxNow and iHealth, for either nasal swabbing (p=1.0) or OP swabbing (p=0.14). With respect to testing method: Nasal AgRDTs tests were more sensitive than OP AgRDTs (all p<0.01). Saliva PCR was significantly more sensitive than OP PCR (p<0.05), and trended towards greater sensitivity than NP PCR, though this did not reach significance (p=0.11). NP and OP PCR testing were not significantly different (Figure 5).

**Figure 2:**
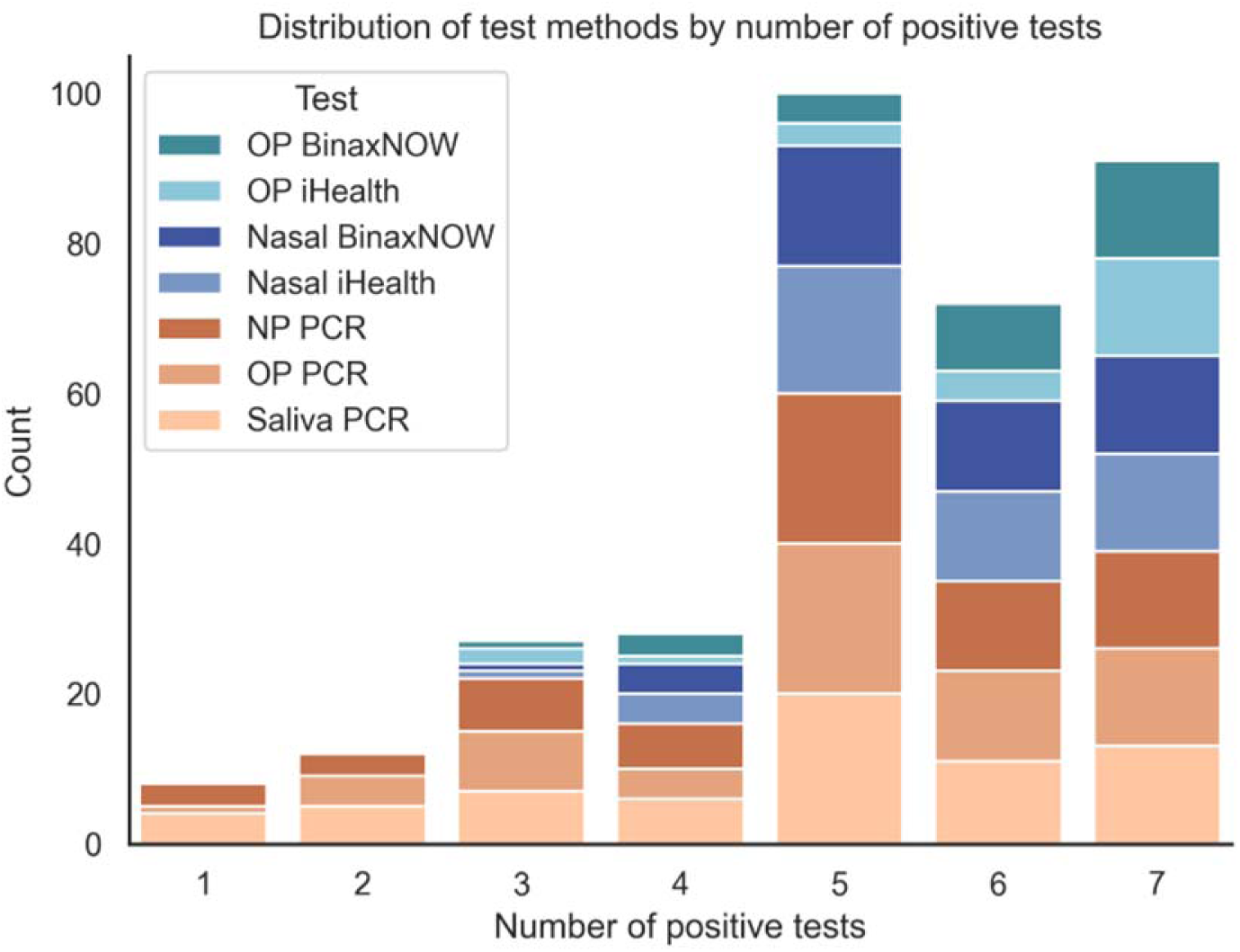
Distribution of test methods by number of positive tests.

**Figure 3:**
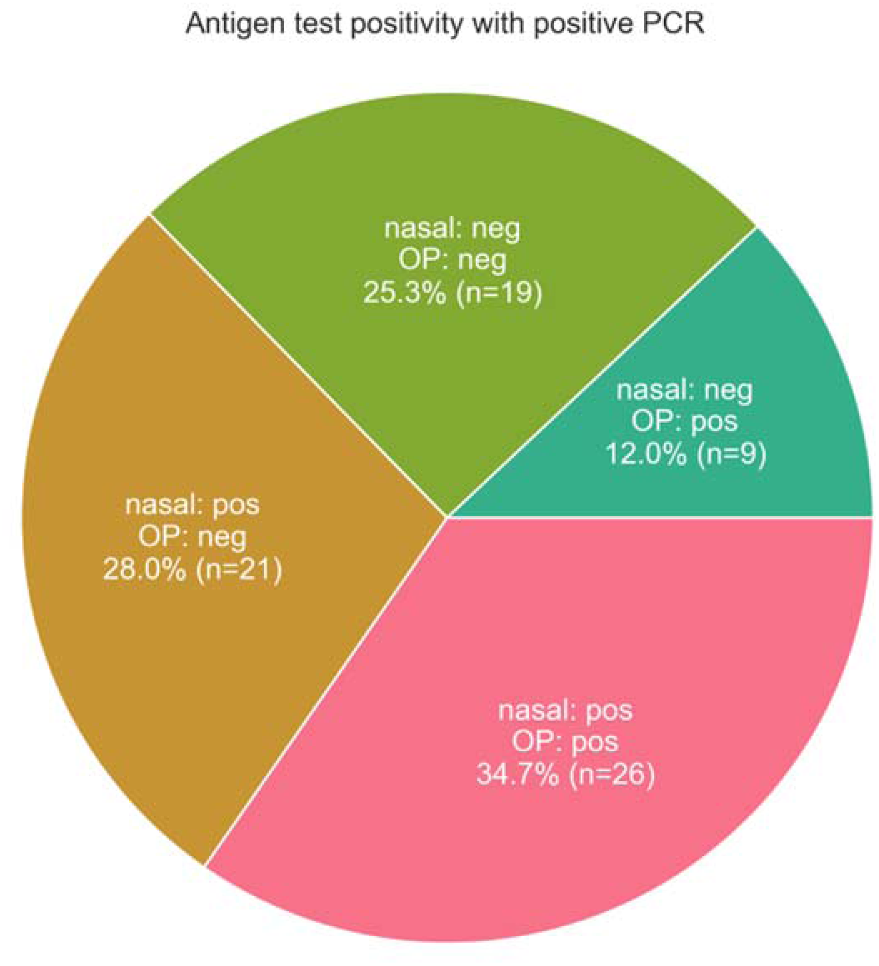
PCR test positivity breakdown in participants with a positive PCR test that 1 or more sites.

**Figure 4:**
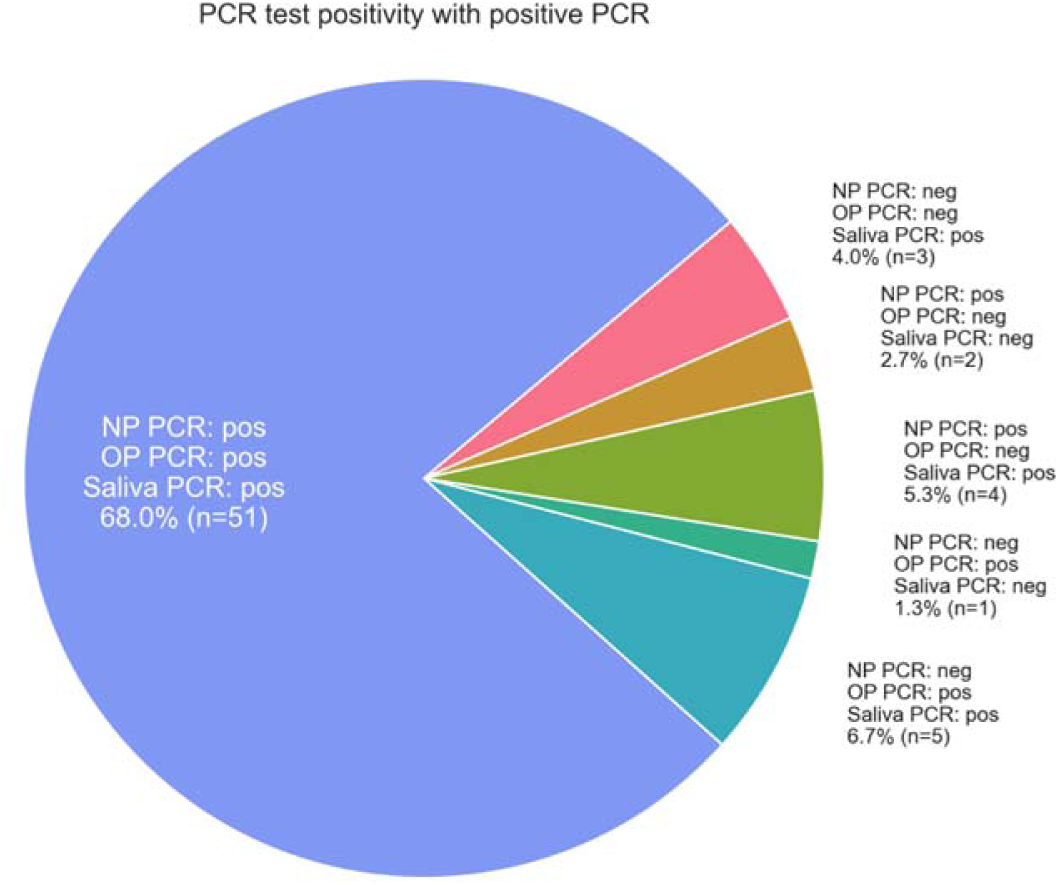
Antigen rapid detection test positivity breakdown in participants with 1 or more positive PCR tests.

**Figure 5:**
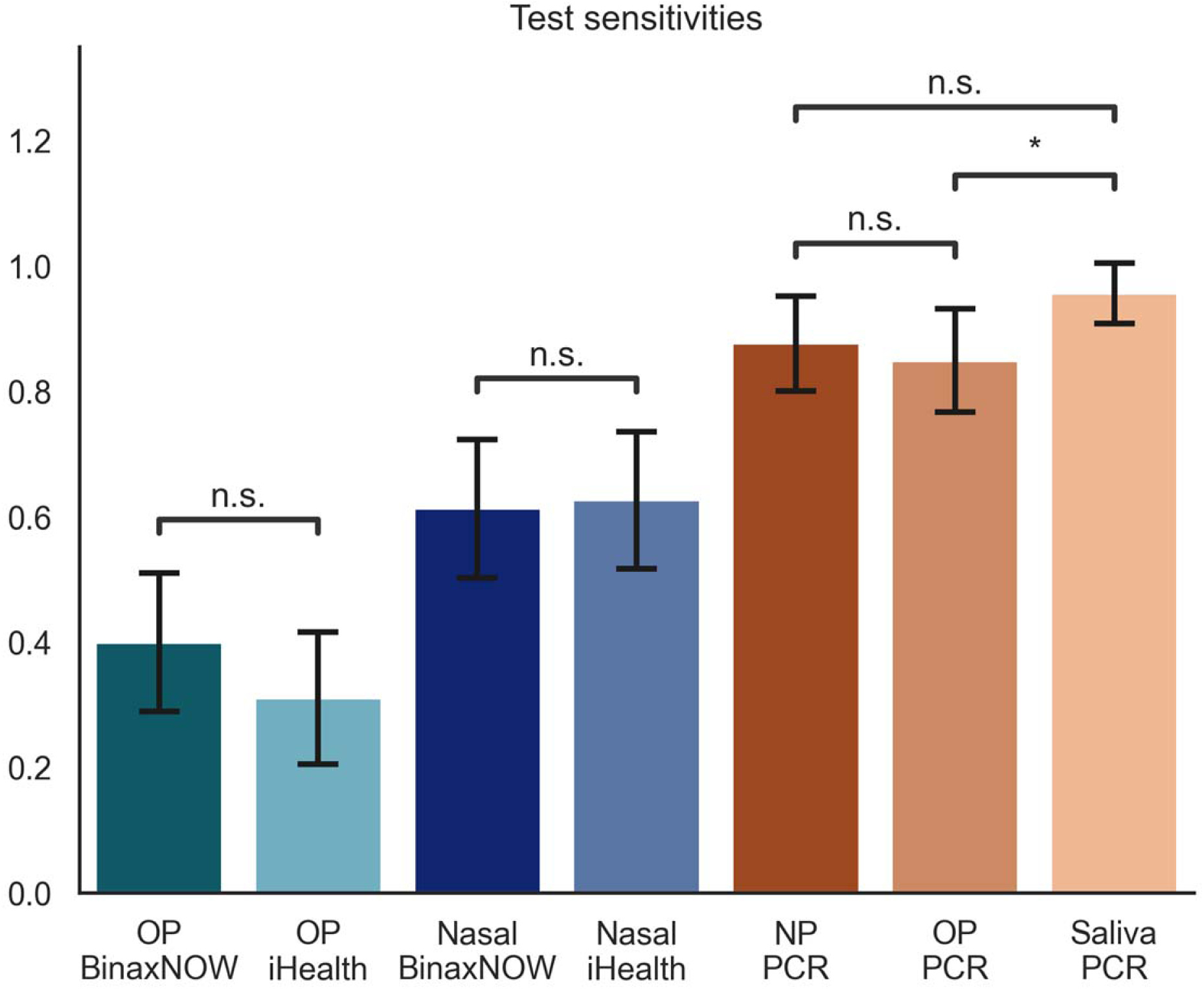
Sensitivities of All Tests. “n.s.” denotes no significant difference between tests. “*” denotes significant difference with p<0.05. All other comparisons not marked are significantly different with p<0.01.

**Table 4:** Sensitivity and Specificity of all tests measured against any positive PCR as gold standard.

| Test | Sensitivity | Specificity |
| --- | --- | --- |
| OP BinaxNOW | 0.400 ± 0.111 | 1.000 ± 0.000 |
| OP iHealth | 0.311 ± 0.105 | 1.000 ± 0.000 |
| Nasal BinaxNOW | 0.613 ± 0.110 | 1.000 ± 0.000 |
| Nasal iHealth | 0.627 ± 0.109 | 1.000 ± 0.000 |
| NP PCR | 0.877 ± 0.075 | 1.000 ± 0.000 |
| OP PCR | 0.849 ± 0.082 | 1.000 ± 0.000 |
| Saliva PCR | 0.957 ± 0.048 | 1.000 ± 0.000 |

*Sequenced Lineages*: 115 samples (including multiple samples from individual participants) were sequenced for Pango Lineage and Nextstrain Clade (Table 5) yielding 8 viral strains by Pango lineage. Ba.1.1 and 21K (Omicron) were the most common, respectively.

**Table 5:** Number of samples in each Pango lineage and Nextstrain clade.

|  | **Pango lineage** |  | **Nextstrain clade** |
| --- | --- | --- | --- |
| **BA.1.1** | 82 | **21K (Omicron)** | 111 |
| **BA.1.15** | 9 | **21L (Omicron)** | 4 |
| **BA.1.20** | 8 |  |  |
| **BA.1.1.18** | 4 |  |  |
| **BA.2** | 4 |  |  |
| **BA.1.17** | 4 |  |  |
| **BA.1** | 2 |  |  |
| **BA.1.17.2** | 2 |  |  |

## Discussion

We compared seven test modalities for the diagnosis of COVID-19 in 325 participants. We found that PCR tests were the most sensitive, nasal AgRDTs were moderately sensitive, and OP AgRDTs were the least sensitive. Prior meta-analyses showed single AgRDTs to have a sensitivity of approximately 70%, and previous studies of BinaxNOW demonstrated a sensitivity of around 65%^5,6^. AgRDT test performance in nasal samples in our study did not differ significantly from these published results. PCR tests had higher sensitivity than all AgRDTs, roughly consistent with previous research on PCR test sensitivity^10^.

Site of testing was also an important factor. Saliva tests were significantly more sensitive than OP tests, but NP tests were not significantly different from saliva or OP tests. This is consistent with prior research on the Omicron variant^14^. Saliva tests are less uncomfortable and less expensive than NP tests^9,15^, which combined with their equal sensitivity to NP tests would appear to make them a preferred test method. One downside to saliva testing is the need for participants to abstain from eating or drinking for 30 minutes prior to testing. During major infection waves such as Omicron, maximizing testing throughput was important, and a 30-minute wait time could be problematic. However, saliva testing can be performed by lay individuals, or even as part of an unsupervised “drop-box” collection system, and could also represent a substantial benefit when testing in the pediatric population.

Use of oropharyngeal swabbing for AgRDTs is a non-standard technique, which in this study yielded lower sensitivity than nasal AgRDTs. Nevertheless, 9 out of 75 positive participants were positive on at least one OP AgRDT, but negative on both nasal AgRDTs, highlighting considerable test performance variability by swabbing site. The reason for this discrepancy is unknown, and could represent variability in individual test kit performance, technical factors during swabbing and/or actual site-specific biological variability in shedding of viral antigens. This observation raises the question as to whether sensitivity during point-of-care or home use of AgRDTs might be optimized by the use of both an oral and a nasal swab during testing. A study by Goodall et al. in 2022 examining the use of OP swabs and combined nasal/throat swabs during the omicron wave found different results, with both NP and OP swabs having 0.645 sensitivity compared to PCR^19^. They also found that combined nasal/throat swabs had an increased sensitivity compared to PCR at 0.887. Important differences between this study and our work include their use of asymptomatic participants, RT-PCR from residual viral media, Panbio brand tests, and sample self-collection. More research is needed to elucidate potential benefits of OP swabs in conjunction with nasal swabs and/or utility of combined nasal/throat swabs.

While early predictive models had found loss of taste and smell, fatigue, cough, and anorexia to be highly correlated with a positive COVID-19 test^20^, in our series Individual symptoms most predictive of a positive test were cough, chills, and fever, with chills and cough being the most significant predictive pair of a positive test. Loss of taste and smell were not commonly reported symptoms in our dataset, and fatigue was quite common but less predictive than many other symptoms. Other studies performed during prevalence of the Omicron variant reported patterns of symptomatology with those observed here^21^, underscoring the need for continued surveillance and clinical research during the evolution of a viral pandemic. Among 115 samples yielding viral genomic sequences, 82 were BA.1.1, and all were within the Omicron family, suggesting most infections in our cohort were due to the omicron variant. A comparative assessment of symptoms and test performance by viral strain was therefore not possible.

This study has several limitations. During data collection, some AgRDTs tests were run at outside ambient seasonal temperatures ranging from 36°F – 60°F. This complies with storage recommendations for both brands (35.6°F and 86°F). However, iHealth manufacturer instructions recommend running the assays between 65-86 °F, and slightly colder temperatures in our “real-world setting” may have affected test performance. Our data set only included one asymptomatic positive individual, who was positive on all three PCR tests negative on all AgRDTs. A 2023 meta-analysis showed decreased sensitivity at 42.6% when AgRDTs were used as screening tools in the general population^4^. AgRDTs therefore may be most valuable for maximizing early detection of COVID-19 in symptomatic individuals but should not be relied on exclusively for all testing purposes.

Conclusion: PCR COVID-19 tests are the most sensitive, and should remain the gold standard for COVID-19 detection. Testing by PCR in saliva is more sensitive than PCR using OP samples, and offers financial, operational, and patient comfort advantages compared to NP tests. AgRDTs are less sensitive than PCR tests. AgRDTs conducted in the standard nasal manner are more sensitive than AgRDTs conducted with oropharnygeal samples. Differences in both test sensitivity and COVID-19 symptom presentation between our data and older, pre-Omicron data reinforce the need for continued assessment of tests and risk stratification tools as pandemics evolve.

## Data Availability

All data produced in the present study are available upon reasonable request to the authors.

## Acknowledgments

We gratefully acknowledge the Occupational Health team, including Kira Loken, Skylar Workinger, Andrea Dayot, Natlie Dutro, and the rest of the occupational health team. We are grateful for the assistance of Oregon National Guard members Austin Krause, Charlie Jedda, and Jack Kelly. We thank the OHSU Molecular Microbiology Lab for support with laboratory testing, the Oregon Health Authority for providing additional rapid antigen tests and medical student researchers Mikaela Siegel, Emily Burney, Ying Yu, and Lynn Bajorek for assistance with recruiting and testing. Lastly, we are especially appreciative of the OHSU workforce members for their extraordinary generosity in participating in this study.

